# Exploring Outcomes and Engagement Patterns of Hypertensive Patients: An Observational Analysis of the Withings Remote Patient Monitoring Program

**DOI:** 10.1101/2024.10.28.24315321

**Authors:** Philomène Letzelter, Benjamin Vittrant, Rachel Tunis

## Abstract

**Background:** Hypertension is a health issue that affects more than one billion people worldwide and can lead to severe health complications. If untreated, hypertensive crises can occur, which are medical emergencies that require immediate care. Remote Patient Monitoring (RPM) offers a transformative approach to managing hypertension by enabling continuous collection and analysis of blood pressure (BP) data. Our study focuses on Withings’ RPM system and analyzes the outcomes and engagement patterns of patients using RPM for hypertension management.

**Methods:** The study comprises 1,873 patients over 6 months who followed an RPM program and used Withings Blood Pressure Monitors. Data was analyzed using Python and R. The analysis examines patients’ systolic and diastolic BP over time as well as the frequency of measurements that patients took using the RPM system.

**Results:** The study reveals that patients who consistently followed the program experienced significant reductions in both systolic and diastolic BP levels. Statistically significant reductions in SBP were observed in users with higher grades of hypertension over six months. The percentage of users experiencing hypertensive crises was reduced from more than 8% to less than 3% over 24 weeks. More frequent engagement with the RPM program was associated with greater BP reductions.

**Conclusion:** These findings highlight the potential of RPM programs in helping patients manage hypertension and minimize hypertensive crises. Future research should go further to understand factors that impact patient outcomes and engagement patterns to improve the effectiveness of RPM programs.

## Introduction

Hypertension, defined as a blood pressure (BP) level of ≥140/90 mmHg by the World Health Organization (WHO), affects approximately 1.28 billion adults globally (1) and accounts for 10.4 million deaths each year (2). In the United States, nearly half of the adult population (119.9 million) is hypertensive (3,4). If untreated, hypertension can lead to severe complications such as diabetes, stroke, chronic kidney disease, cardiovascular disease, and more. Even more troublingly, hypertension is often referred to as the “silent killer”, as it is often asymptomatic and, despite posing significant health risks, many hypertensive patients may be unaware of its presence. With treatment (i.e., medication to lower BP) and lifestyle changes, hypertension can be managed, yet low adherence to antihypertensive medication is a significant issue, with a nonadherence rate of 31% among insured adults (5). Still, even achieving small reductions in BP can make a huge difference for patients’ overall health. Research has found, for example, that even a 5 mmHg reduction in systolic BP reduces the risk that a patient will experience a major cardiovascular event by 10 percent (6).

As such, to ensure proper management of hypertension, continuous BP monitoring is crucial for hypertensive patients and those at risk of developing hypertension. Increasingly, digital BP monitors coupled with Remote Patient Monitoring (RPM) programs allow patients and their healthcare providers to remotely monitor BP readings taken at home with the purpose of improving treatment adherence and preserving limited resources. Such digital disease management tools have been praised by health and technology stakeholders for their potential to improve patients’ health outcomes (7), and some studies of RPM platforms have demonstrated that participants experience statistically significant decreases in BP (8–10). For example, Persell et al. found that among their study participants using RPM, systolic BP was lower and BP control was better as compared with the control group, with statistically significant differences persisting for at least 18 months (9,11). Leo et al. (8) performed a systematic review of 96 studies and also found that RPM led to statistically significant decreases in both systolic and diastolic BP (SBP and DBP).

Other studies, however, have noted the “limited evidence” that exists to substantiate the claims of RPM and found these programs to have a non-significant impact on clinical outcomes (12). While some clinical trials did not find statistically significant differences in BP between an RPM group and a control group overall, significant reductions were still observed in specific sub-groups. For example, Kim et al. (13) observed a significant decrease in systolic BP among patients aged 55 or older who used RPM for BP management. This suggests that more research is needed to understand not only which groups of patients might benefit most from RPM, but how their level of engagement (i.e., the frequency and duration of their RPM use) impacts their outcomes. Some research has suggested that increased engagement in digital health management could be linked to improved clinical outcomes (14), but as previous studies have pointed out, published data on the impact of RPM systems on both clinical outcomes and engagement are limited (10,12,14).

This study aims to fill this research gap by analyzing data on patient outcomes and engagement from the Withings RPM program (15) over a 24-week period. The Withings RPM system allows patients to take their BP readings at home and enables healthcare professionals to remotely access patient health records and monitor health metrics. This facilitates the creation of customized treatment plans, alerts for out-of-range data, and timely patient engagement to improve medication adherence and health outcomes. By analyzing both clinical outcomes and engagement patterns of patients using the Withings RPM system over 24 weeks, we contribute to the field’s understanding of the impact and potential of RPM for various groups of hypertensive patients. More specifically, our findings provide insights into the characteristics and engagement patterns of patients who experience a significant decrease in BP over the course of their engagement with Withings RPM. Finally, our study points the way for future research to undertake even more detailed analyses to better understand trends in outcomes and engagement data and ultimately maximize the effectiveness of RPM programs for hypertension patients broadly.

## Methods

### Dataset

All users of the RPM program who had at least one Withings BP Monitor as of June 2024 were included in our dataset. Inclusion criteria required patients to be 18 years or older and to have at least 24 weeks between their first and their most recent measure. This led to the selection of 1,877 users, predominantly from the United States (99.6%). The data was anonymized and aggregated to ensure HIPAA compliance. Descriptive statistics for the sample are shown in Table 1.

**Table 1:**
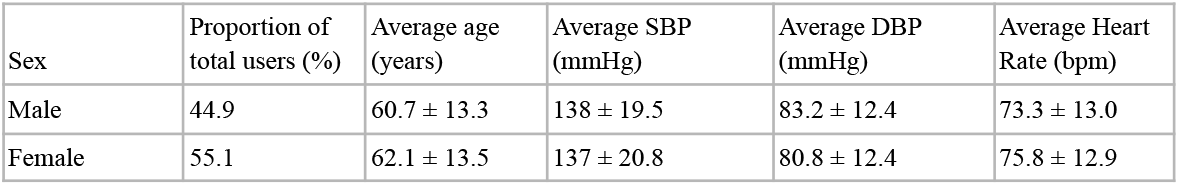
Descriptive statistics of sample used in analysis.

| Sex | Proportion of total users (%) | Average age (years) | Average SBP (mmHg) | Average DBP (mmHg) | Average Heart Rate (bpm) |
| --- | --- | --- | --- | --- | --- |
| Male | 44.9 | 60.7 ± 13.3 | 138 ± 19.5 | 83.2 ± 12.4 | 73.3 ± 13.0 |
| Female | 55.1 | 62.1 ± 13.5 | 137 ± 20.8 | 80.8 ± 12.4 | 75.8 ± 12.9 |

### RPM Program

Participants in this study took their in-home BP measurements using the Withings BPM Connect or BPM Connect Pro devices. These clinically validated devices measure BP and heart rate using the cuff oscillometric method, with a pressure range of 0 - 285 mmHg (diastolic range 40 - 130 mmHg, systolic range 60 - 230 mmHg), with an accuracy of ± 3 mmHg and a pulse rate range of 40 - 180 beats/min (16–18). Patients enrolled in the Withings RPM program are regularly monitored by clinical staff, with a clear protocol for responding to out-of-target readings and escalating potential emergency situations to ensure timely medical intervention. Withings provides both the measuring devices and the platform to facilitate the monitoring of patients’ vitals and care management. Program specifics, including patient eligibility, objectives, protocols and consent forms are specific to and handled independently by each care team (15).

### Classification of BP Levels

BP levels were categorized following American Heart Association (AHA) guidelines (4,19) into Normal, Elevated, Grade 1 and Grade 2 Hypertension. We also included an additional category, Grade 3 Hypertension, due to the significant proportion of users with BP ≥ 160/100 mmHg (12.8%).

Therefore, we use the following thresholds for classification in this analysis:

- Normal : SBP < 120mmHg and DBP < 80 mmHg
- Elevated : SBP 120 - 129 mmHg and DBP < 80 mmHg
- Grade 1 : SBP 130 - 139 mmHg or DBP 80 - 89 mmHg
- Grade 2 : SBP 140 - 159 mmHg or DBP ≥ 90 mmHg
- Grade 3 : SBP ≥ 160 mmHg or DBP ≥ 100 mmHg

Further, a critical risk for hypertensive patients is the occurrence of hypertensive crises, defined by the AHA as BP ≥ 180/120 mmHg (20). These crises are considered medical emergencies due to the important risk of organ damage, and they require immediate medical care. Therefore, hypertensive crises were also analyzed in this study.

### Data Analysis

Data analysis was performed using Python v3.9.6 (21) and R version 4.4.1 (June 2024) (22). Plots were created using Rstudio v2024.04.0 build 735 (23) with R library ggplot 2 (24), and statistical analyses were conducted using the ttest_1samp function from the scipy.stats module (25) in Python. Paired t-tests were used to compare differences in users’ mean SBP between weeks 1 and weeks 6, 12, and 24. Anderson-Darling tests were performed to ensure that the data met normality assumptions (26). All users were compared by ‘relative week’ based on the date of the first BP measurement taken, to account for differences in program dates and durations. User engagement was assessed by analyzing the frequency of blood pressure measurements users took per week, described further in the next section. Covariates such as age, sex, and BMI were not adjusted for in this study.

### Ethical considerations

The data used in this analysis is secondary data that was taken throughout the regular course of care by Withings partners using the RPM program. All data has been aggregated and anonymized in this analysis to comply with HIPAA as well as European data protection regulations (27,28). Since the data were aggregated and contained no personal identifiers or protected health information, the involvement of an ethical committee was not required.

## Results

### BP outcomes

Within the first week of the program (relative to each user), 13.6% of users had normal BP, 12.4% had elevated BP, 28% were classified as Grade 1, 33.3% (647 users) as Grade 2, and 12.8% (225 users) as Grade 3. Among the Grade 3 category, 8.3% (21 users) had a BP of ≥180/120 mmHg, which is classified as a hypertensive crisis and presents a life-threatening risk.

Our results indicate statistically significant reductions in SBP and DBP over 24 weeks for users with more severe hypertension (Grade 2 and Grade 3). The mean reduction in SBP was 23 mmHg for Grade 3 and 8 mmHg for Grade 2 after 24 weeks (Figure 1), both statistically significant (p < 0.001). Conversely, users with normal and elevated BP experienced small increases in BP over 24 weeks, also both statistically significant (p < 0.001) (Table 2). There was no significant increase or decrease for users with Grade 1 hypertension.

**Table 2:** Differences in SBP by week (as compared with week 1), by BP category.

|  | Mean Difference in SBP (mmHg) | 95% CI Lower | 95% CI Upper | Significance (P-value) |
| --- | --- | --- | --- | --- |
| **Week 6** |  |  |  |  |
| Normal | 5.95 | 4.18 | 7.72 | **<0.001** |
| Elevated | 1.39 | -0.10 | 2.89 | 0.07 |
| Grade 1 Hypertension | -0.39 | -1.34 | 0.56 | 0.42 |
| Grade 2 Hypertension | -5.20 | -6.26 | -4.13 | **<0.001** |
| Grade 3 Hypertension | -15.42 | -17.69 | -13.15 | **<0.001** |
| **Week 12** |  |  |  |  |
| Normal | 6.97 | 5.34 | 8.59 | **<0.001** |
| Elevated | 3.42 | 1.79 | 5.06 | **<0.001** |
| Grade 1 Hypertension | -0.54 | -1.68 | 0.61 | 0.36 |
| Grade 2 Hypertension | -6.17 | -7.32 | -5.01 | **<0.001** |
| Grade 3 Hypertension | -18.04 | -20.55 | -15.53 | **<0.001** |
| **Week 24** |  |  |  |  |
| Normal | 11.03 | 8.90 | 13.16 | **<0.001** |
| Elevated | 5.57 | 3.77 | 7.37 | **<0.001** |
| Grade 1 Hypertension | 0.55 | -0.58 | 1.67 | 0.34 |
| Grade 2 Hypertension | -7.92 | -9.23 | -6.61 | **<0.001** |
| Grade 3 Hypertension | -22.96 | -25.85 | -20.08 | **<0.001** |

**Figure 1:**
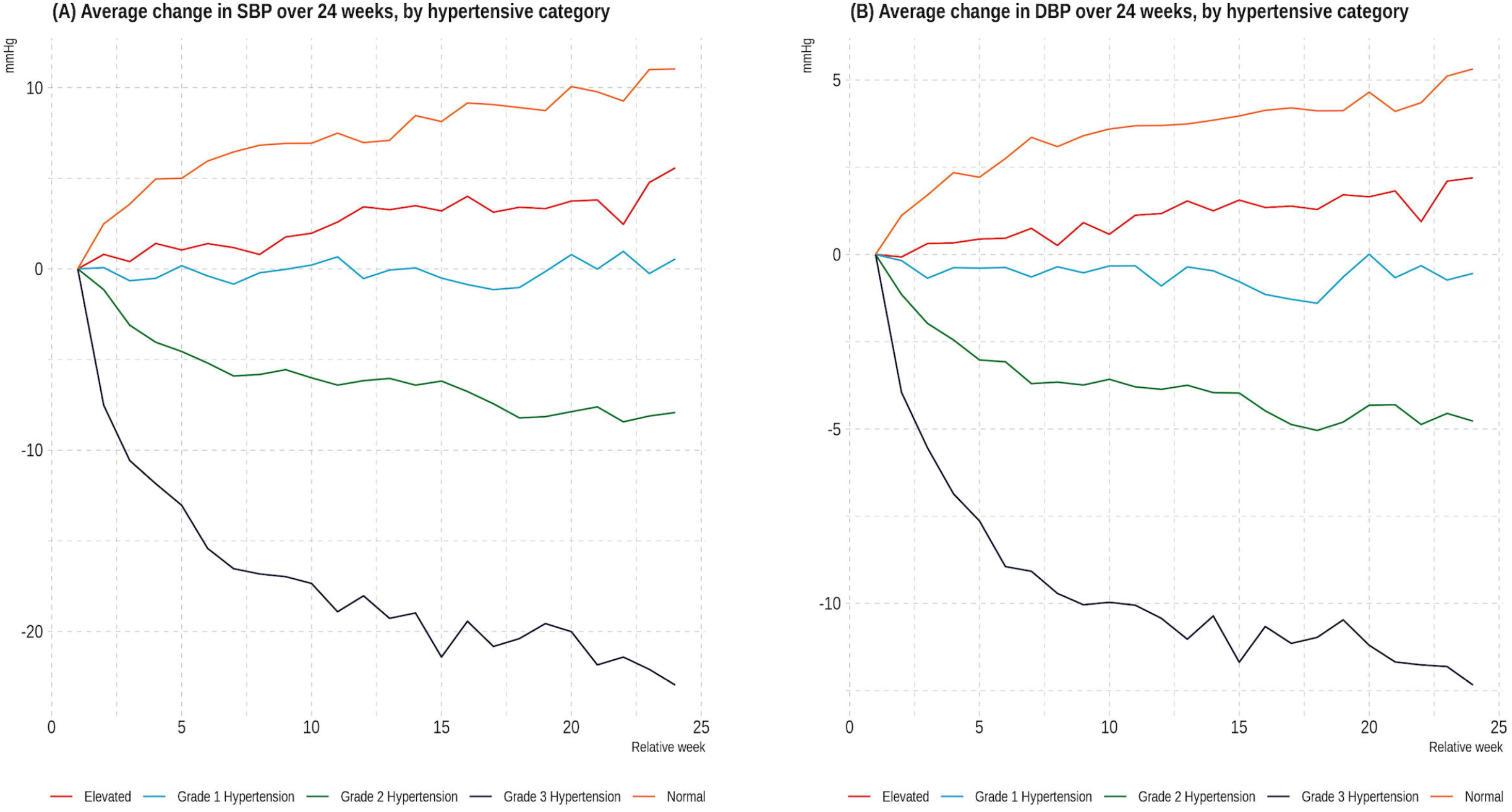
(A) Average change in SBP over 24 weeks, by hypertensive category; (B) Average change in DBP over 24 weeks, by hypertensive category

As previously stated, 8.3% of users had at least one BP reading ≥180/20 mmHg in their first week of measurements (indicating a hypertensive crisis). Of these users, 1% consistently experienced a mean BP of ≥180/20 during the first week (i.e., in more than three measurements). Additionally, among the entire cohort, a total of 673 users (28.6%) experienced a hypertensive crisis at some point over the course of 24 weeks. This is an abnormally high proportion compared to literature indicating that approximately 1-2% of hypertensive patients experience a hypertensive crisis (29–31). Therefore, it is a major point of interest to quantify the reduction in the percentage of users experiencing crises over time through regular use of RPM, as presented in Figure 2. Our data show a high initial prevalence of hypertensive crises, with a significant reduction over time. As Figure 2 indicates, there was also a gradual decline in the number of patients recording blood pressure measurements over the 24-week period, which is typical after the introduction of a technological intervention. Nevertheless, the proportion of active patients experiencing hypertensive crises decreased significantly, from 8.3% in week 1 to less than 3% by week 24.

**Figure 2:**
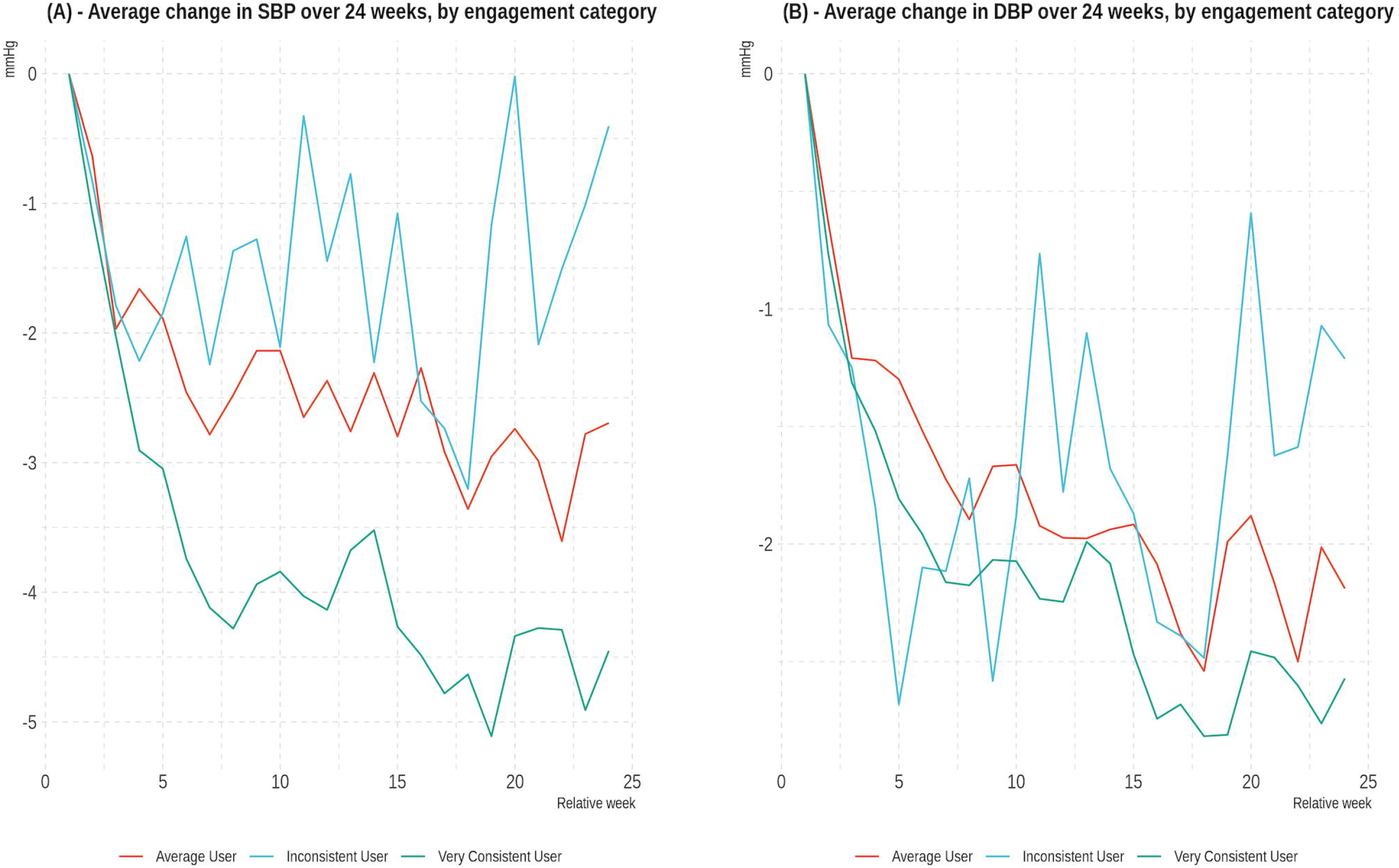
Percentage of users experiencing hypertensive crisis per week

### Engagement Analysis

Users recorded a median of 5 measures per week and 18 measures per month. The data indicates high consistency in user activity, with users taking measures in a median of 92.5% of the weeks, and 100% of the months during their engagement period, spanning the period between their initial and their most recent measurement. Engagement was categorized based on measurement frequency, which was segmented by quartiles:

- Very Consistent users: More than 5.5 measurements per week (Q3)
- Consistent users: Between 3 and 5.5 measurements per week (IQR)
- Inconsistent users: Fewer than 3 measurements per week (Q1)

Figure 3 illustrates the mean difference in BP at each relative week rank compared to week 1, averaged across all users, and segmented by engagement category. Paired t-tests were also conducted to examine the reduction in SBP for users in each engagement category between week 1 and weeks 6, 12, and 24 (Table 3). At each of these time points, statistically significant reductions were observed for users in the “average” and “very consistent” engagement categories, but not the “inconsistent” category (p < 0.001). The reductions were the greatest for very consistent users, who saw a mean difference of 4.45 mmHg over 24 weeks. As Table 4 shows, the frequency of measures was largely not correlated with users’ initial BP levels, with the exception of users with Grade 3 hypertension, who were less engaged overall.

**Table 3:** Differences in SBP by week (as compared with week 1), by engagement category.

|  | Mean Difference<br>in SBP (mmHg) | 95% CI Lower | 95% CI Upper | Significance<br>(P-Value) |
| --- | --- | --- | --- | --- |
| **Week 6** |  |  |  |  |
| Inconsistent | -1.26 | -3.21 | 0.70 | 0.207 |
| Average | -2.46 | -3.36 | -1.56 | **<0.001** |
| Very consistent | -3.74 | -4.84 | -2.64 | **<0.001** |
| **Week 12** |  |  |  |  |
| Inconsistent | -1.45 | -3.69 | 0.80 | 0.206 |
| Average | -2.37 | -3.38 | -1.36 | **<0.001** |
| Very consistent | -4.14 | -5.35 | -2.93 | **<0.001** |
| **Week 24** |  |  |  |  |
| Inconsistent | -0.40 | -2.93 | 2.12 | 0.753 |
| Average | -2.69 | -3.90 | -1.49 | **<0.001** |
| Very consistent | -4.45 | -5.98 | -2.92 | **<0.001** |

**Table 4:** Frequency of user engagement by blood pressure category.

| BP category | Median total measures | Median measures/ month | Median measures/ week |
| --- | --- | --- | --- |
| Normal | 140.5 | 19.0 | 5.0 |
| Elevated | 152.5 | 19.0 | 5.0 |
| Grade 1 | 141.0 | 17.0 | 5.0 |
| Grade 2 | 140.0 | 17.0 | 5.0 |
| Grade 3 | 109.0 | 15.0 | 4.0 |

**Figure 3:**
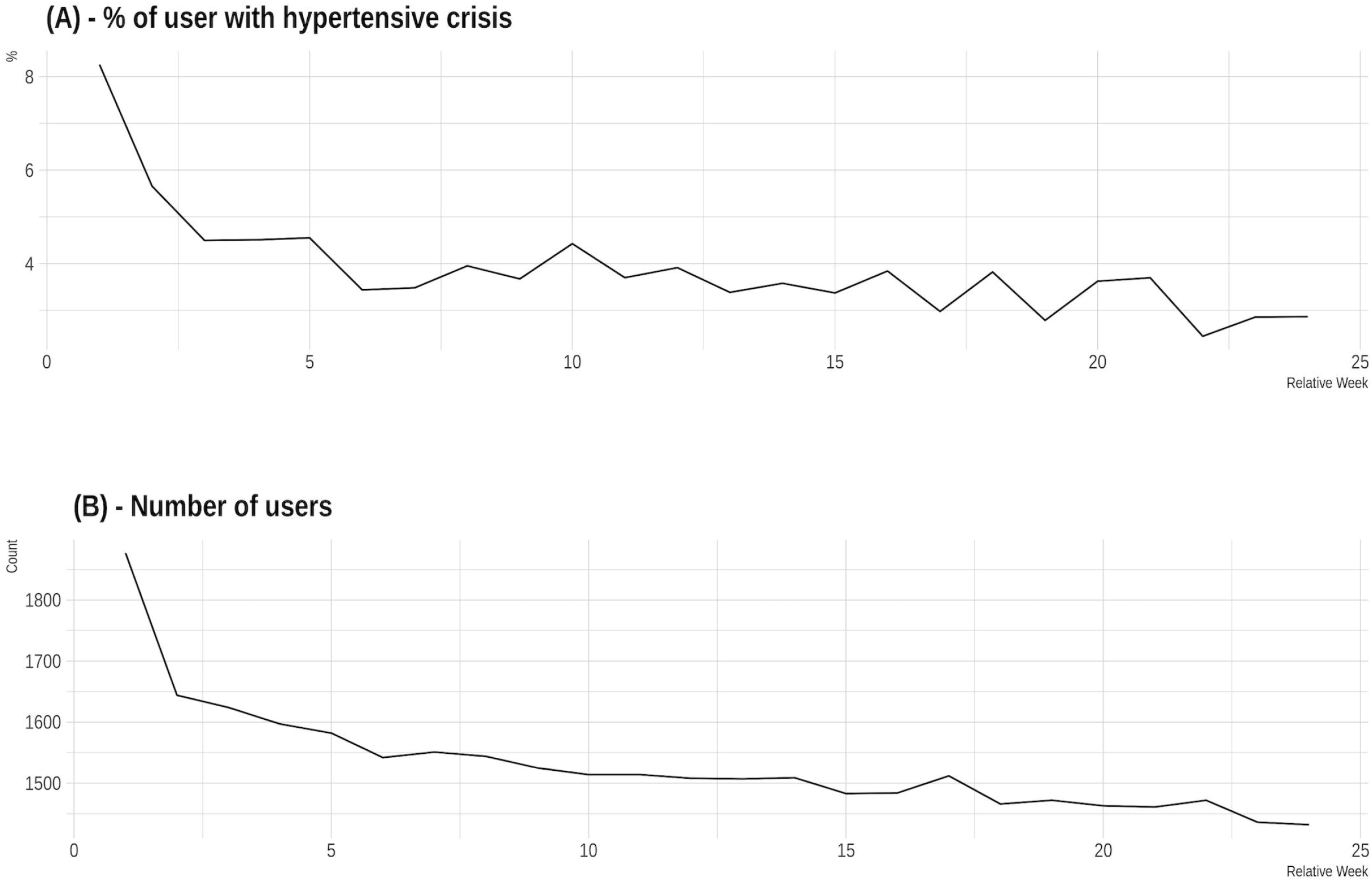
Average change in BP over 24 weeks, by engagement category

## Discussion

This analysis showed significant reductions in BP for users with more severe hypertension (in Grades 2 and 3) over the course of 24 weeks of RPM. This was anticipated since the data were derived from a clinical program for hypertension focused on patient BP monitoring. The fact that these reductions were seen in users with higher stages of hypertension rather than other categories could be due in part to the fact that the care team prioritizes responding to hypertensive crises, or to the fact that patients with a lower grade of hypertension might be dealing with a primary medical indication not directly related to hypertension (e.g., pregnancy, atrial fibrillation), and so competing priorities are at play. Other studies have found, similarly, that patients at more advanced stages of a chronic condition (for example, diabetic patients who have a higher HbA1C) are more likely to engage frequently and see better outcomes when using digital health management tools (32,33). Kim et al. (13) found, relatedly, that older patients were more likely to see a significant decrease in BP, a finding which may align with the strong potential of RPM for higher-risk patients. It is promising that patients at higher risk seem to be seeing the most significant results from RPM; this finding highlights the potential of remote behavioral programs to prevent worsening BP and should validate efforts to expand access to RPM and other digital health solutions for high-risk patients. However, it also calls for future research to better understand the engagement patterns and ideal circumstances for lower-grade/ lower-risk patients to reach their BP targets.

Our study also analyzed engagement patterns, finding that more regular, consistent users saw a greater reduction in SBP than inconsistent users. Frazier et al. (10) also studied the correlation of engagement patterns with clinical outcomes and found that while BP did decrease the more frequently patients took measurements, the reduction was not significant, and no minimum number of measurements predicted the failure of RPM to lower BP. Our findings contrast slightly, indicating that more frequent measurements through RPM are linked to greater reductions in BP. Given that the number of measurements taken per month is part of the requirement for reimbursement of RPM programs by CMS, this is an important area for researchers to continue to assess (34).

We also note that our dataset had a particularly high proportion of users experiencing hypertensive crises. While the strong and significant reduction that we observed in patients experiencing hypertensive crises aligns with our finding that patients with higher grades of hypertension will see more significant reductions using RPM, we cannot be sure of the underlying explanation for such a high proportion of users experiencing a hypertensive crisis in the first place. It is possible that RPM generally facilitates the detection of hypertensive crisis events that would not otherwise be captured in clinical settings, but more research using real-life datasets is needed to provide more precise insights into these poorly understood events.

## Limitations

As with all retrospective observational studies, we are limited by potential bias in the dataset, and we lack a great deal of contextual data that may have helped explain the data. Our analysis did not adjust for covariates to assess other potential explanatory factors associated with BP reductions, which will be essential for future research to build on our results. We also did not have data available to us on many relevant, potentially confounding medical variables. For example, medication adherence is a crucial factor influencing BP outcomes that is frequently used as a way to deliver and assess digital interventions like RPM programs (35,36), and which we were not able to analyze or assess in this study. Similarly, data giving a more comprehensive picture of patients’ lifestyle behaviors, comorbidities, interactions with their care team, and so forth would undoubtedly help paint a clearer picture of the impacts of RPM on different patient groups.

## Conclusion

In this study, we observed that the Withings RPM program has a significant beneficial effect on patients’ BP, and more frequent measurements appear to contribute to this effect. These findings suggest the benefits of RPM programs broadly in managing hypertension and minimizing hypertensive crises. However, further research is needed to identify individual-level factors influencing patient engagement and the overall effectiveness of RPM programs, recognizing that these programs may be more effective for certain patient subpopulations.

## Data Availability

All data produced in the present study are available upon reasonable request to the authors.

## Conflict of interest

Withings is the manufacturer and developer of the Blood Pressure Monitors used and this study was conducted by Withings researchers. While the publication does not intend to make any claims about the performance of Withings devices, the authors declare a potential conflict of interest, as the findings of this study may have implications for the acceptance and use of the company’s products and services.

## CRediT statement

**P.L**.: Conceptualization, Methodology, Software, Formal Analysis, Writing—Original Draft; **B.V**.: Supervision, Visualization, Writing—Original Draft, Validation; **R.T**.: Investigation, Writing– Original Draft, Validation

## Funding

This work was entirely supported by Withings.

## Acknowledgments

We’d like to thank all the Withings collaborators that work on our products from the hardware to the software allowing us to perform this research and observations.

## Notes

### Competing Interest Statement

Withings is the manufacturer and developer of the Blood Pressure Monitor used and this study was conducted by Withings researchers. Even if the publication does not make any explicit claims about the performance of Withings devices, the authors declare a potential conflict of interest, as the findings of this study may have implications for the acceptance and use of the companys products and services.

### Author Declarations

The data used in this analysis is secondary data that was taken throughout the regular course of care by Withings partners using the RPM program. All data has been aggregated and anonymized in this analysis to comply with HIPAA as well as European data protection regulations (27,28). Since the data were aggregated and contained no personal identifiers or protected health information, the involvement of an ethical committee was not required. The use of data in this study was overseen by the Withings Data Protection Officer to ensure adherence to privacy protection standards.

